# Transcutaneous Cervical Vagal Nerve Stimulation Reduces Sympathetic Responses to Stress in Posttraumatic Stress Disorder

**DOI:** 10.1101/2020.02.10.20021626

**Authors:** Nil Z. Gurel, Matthew T. Wittbrodt, Hewon Jung, Md. Mobashir H. Shandhi, Emily G. Driggers, Stacy L. Ladd, Minxuan Huang, Yi-An Ko, Lucy Shallenberger, Joy Beckwith, Jonathon A. Nye, Bradley D. Pearce, Viola Vaccarino, Amit J. Shah, Omer T. Inan, J. Douglas Bremner

## Abstract

**Objective:** Exacerbated autonomic responses to acute stress are prevalent in posttraumatic stress disorder (PTSD). The purpose of this study was to assess the effects of transcutaneous cervical VNS (tcVNS) on autonomic responses to acute stress in patients with PTSD. The authors hypothesized tcVNS would reduce the sympathetic response to stress compared to a sham device.

**Methods:** Using a randomized double-blind approach, we studied the effects of tcVNS on physiological responses to stress in patients with PTSD (n=25) using noninvasive sensing modalities. Participants received either sham or active tcVNS after exposure to acute personalized traumatic script stress and mental stress (public speech, mental arithmetic) over a three-day protocol. Physiological parameters related to sympathetic responses to stress were investigated.

**Results:** Relative to sham, tcVNS paired to traumatic script stress decreased sympathetic function as measured by: decreased heart rate (adjusted ß=-5.7%; 95% CI: ±3.6%, effect size d=0.43, p<0.01), increased photoplethysmogram amplitude (peripheral vasodilation) (30.8%; ±28%, 0.29, p<0.05), and increased pulse arrival time (vascular function) (6.3%; ±1.9%, 0.57, p<0.0001). Similar (p < 0.05) autonomic, cardiovascular, and vascular effects were observed when tcVNS was applied after mental stress or without acute stress.

**Conclusion:** tcVNS attenuates sympathetic arousal associated with stress related to traumatic memories as well as mental stress in patients with PTSD, with effects persisting throughout multiple traumatic stress and stimulation testing days. These findings show that tcVNS has beneficial effects on the underlying neurophysiology of PTSD. Such autonomic metrics may also be evaluated in daily life settings in tandem with tcVNS therapy to provide closed-loop delivery and measure efficacy.

ClinicalTrials.gov Registration # NCT02992899

Highlights
- We studied the effects of tcVNS on physiological responses to stress in patients posttraumatic stress disorder (PTSD).
- tcVNS modulates physiologic reactivity to traumatic and mental stress in PTSD, and modulates autonomic tone when applied without acute stress.
- Repeated tcVNS enhances resilience in the face of repeated stress in PTSD as quantified by peripheral autonomic measures which potentially could serve as real-time measures to evaluate the therapy response in longitudinal settings.

## Introduction

Posttraumatic stress disorder (PTSD) has a lifetime prevalence of 8% and is associated with considerable morbidity, loss of productivity, and treatment costs (1). Less than half of patients with PTSD seek or receive treatment, with existing treatment options exhibiting high (24%) dropout rates due to insufficient time with the mental health professional, treatment ineffectiveness, work interference, personal problems, or discomfort (2). Cognitive behavioral therapy with prolonged exposure is an effective method to improve the PTSD symptoms in some patients, but requires considerable expertise, time, and resources (3, 4). In addition, psychiatrists may hesitate to employ exposure therapies due to concerns about decompensation, discomfort in using exposure, and patients’ reluctance regarding re-exposure to traumatic reminders (5, 6). Pharmacological treatments represent another standard of treatment although questions persist regarding their efficacy (7–9). New, evidence-based treatments that align well with the clinical needs of those with PTSD are needed (10–12).

In patients with PTSD, exposure to events or stress—particularly those with salient characteristics related to previously experienced trauma—can elicit symptoms such as hyperarousal, intrusive thoughts, avoidance behaviors, and dissociation (13). This adverse response can lead to elevated inflammatory marker concentrations, impaired autonomic modulation, memory deficits, changes in brain morphology, and increased neural reactivity in emotion-specific brain areas (13–22). Vagus nerve stimulation (VNS) is a potential treatment method for PTSD as it modulates sympathetic tone and related cardiovascular reactivity (23), along with enhancing fear extinction in rodents trained with conditioned fear paradigm (24, 25); however, it is limited by the cost and inconvenience of surgical procedures as revealed by the high rates of non-compliance in multi-year VNS studies due to the biological and psychological effects of the surgical implantation (26, 27). Recent advances in noninvasive neuromodulation technologies are promising for widespread use of VNS.

Transcutaneous VNS devices noninvasively target vagal projections in the ear (auricular branch of the vagus) and neck (cervical branch in the carotid sheath). Auricular tVNS (taVNS) devices modulate central and peripheral physiology, as observed with the monitoring of peripheral physiological parameters (28–30), inflammatory cytokines (31–33), hormonal indices (34), brain imaging (35–38), and a case study comparing VNS implant and taVNS for an epilepsy patient (39). taVNS also has been shown to ameliorate tinnitus (40, 41), atrial fibrillation (33), episodic migraine (37), seizure frequency (42), cluster headache (43), and major depression (44, 45), as well as improving vagal tone, deactivation of limbic and temporal brain structures, and mood enhancement in healthy populations and patients with PTSD and mild traumatic brain injury (28, 30, 46, 47). Fewer studies have looked at cervical tVNS (tcVNS), although they have been shown to reliably activate vagal nerve fibers (48–50), produce anti-inflammatory effects (51–53), reduce neural and physiologic responses to noxious thermal stimuli (54) with possible clinical utility in migraine and trigeminal allodyna (55, 56). We have recently explored pairing of tcVNS with personalized traumatic script stress and non-personalized mental stress (public speaking, mental arithmetic) in healthy individuals with a history of exposure to psychological trauma without the current diagnosis of PTSD, and shown a reduction in cardiovascular reactivity and peripheral sympathetic activity both for personalized traumatic stress and neutral mental stress as well as in the absence of stress exposure (57). In this study, we examined the physiological effects of tcVNS in patients with PTSD to traumatic and mental stress. The physiological measurements included the collection of electrocardiography (ECG), seismocardiography (SCG), photoplethysmography (PPG), respiration (RSP), electrodermal activity (EDA), and blood pressure (BP). We hypothesized that tcVNS (compared to sham) would attenuate the physiological responses to stress in PTSD.

## Materials and Methods

### Participants & Assessments

The study was approved by the Institutional Review Boards of Georgia Institute of Technology, Emory University, SPAWAR Systems Center Pacific, and the Department of Navy Human Research Protection Program and was conducted at the Emory University School of Medicine between May 2017 and October 2019. It should be noted that the study registration (ClinicalTrials.gov # NCT02992899) included autonomic peripheral measures examined in this study, however these were not listed as primary or secondary outcomes, rather the outcomes focused on previously examined blood biomarkers by our group in PTSD populations (58). The physiological reactivity measures examined in this study are exploratory research questions that align well with the effects of tcVNS. Participants in the current study aged between 18-70 years with current PTSD as determined by the Structured Interview for Diagnostic and Statistical Manual of Mental Disorders (SCID) (59) and provided written, informed consent. Exclusion criteria were: pregnancy; meningitis; traumatic brain injury; neurological disorder; organic mental disorder; history of loss of consciousness greater than one minute; alcohol abuse or substance abuse based on the SCID within the past 12 months; current or lifetime history of schizophrenia, schizoaffective disorder, or bulimia, based on the SCID; a history of serious medical or neurological illness, such as cardiovascular, gastrointestinal, hepatic, renal, neurologic or other systemic illness; evidence of a major medical or neurological illness on physical examination or as a result of laboratory studies; active implantable device (i.e. pacemaker); carotid atherosclerosis; cervical vagotomy. The Clinician Administered PTSD Scale (CAPS) was administered to establish current PTSD diagnosis and quantitate severity of symptoms (60). Although the results presented here focused on participants with PTSD, the overall study involved both PTSD and non-PTSD traumatized controls and the recruitments were made regardless of the PTSD status, depending on the availability of the participant pool. The physiological reactivity results regarding the non-PTSD population were published in (57, 61). Within the main (PTSD and non-PTSD) study, among 129 who were screened for eligibility (See Figure S1 for Consolidated Standards of Reporting Trials (CONSORT) diagram), 69 excluded due to lack of consent or not meeting the inclusion criteria. The remaining 60 were randomized to active stimulation or sham. Among 60 randomized participants, 35 excluded due to the following reasons: participant did not come or protocol did not continue due to high-resolution positron emission tomography (HR-PET) scanner malfunction used in the study (explained in study protocol below) or the participant did not have PTSD. Among the remaining 51, 26 participants were non-PTSD traumatized controls and 25 were patients with PTSD. In the current study, data from 25 participants with PTSD were analyzed. Table S1 presents the demographics data. The mean age was 35 (±13 SD) with 19 females. The active group participants (n=13) had a mean age of 33 (±12 SD) and included 12 females; sham group participants (n=12) had a mean age of 38 (±13 SD), with seven females. SCID was used to evaluate for possible co-morbid psychiatric diagnosis. In this sample, 13 (52%) met criteria for major depressive disorder (MDD, 6 current, 7 past), eight (32%) for generalized anxiety disorder, four (16%) for panic disorder, two (8%) for social phobia, two (8%) for current obsessive compulsive disorder, one (4%) for agoraphobia without panic disorder, one (4%) for body dysmorphia, three (12%) for past alcohol abuse or dependence, and one (4%) for a past substance induced anxiety disorder.

### Study Protocol

The three-day protocol is summarized in Figure S2. Participants were instructed to withhold any stimulant (i.e. coffee) throughout the entire protocol. Participants provided their traumatic experiences in written form, later, personalized traumatic stress scripts were prepared for each participant. On day 1, participants were prepared with noninvasive sensing modalities (explained in physiological monitoring section), dedicated neuromodulation devices, and a headphone by the researchers. The protocol started when they lay down in an HR-PET scanner bed (head inside the scanner) for 14 HR-PET scans, each took approximately eight minutes. Before the scans started, baseline physiological data were collected in the same posture. In the first two scans, “neutral” pleasant scenery recordings (for imaging purposes, without stimulation) were delivered audibly. In scans three and four, traumatic stress recordings were delivered immediately followed by stimulation. In scans five and six, stimulation was applied in the absence any acute stressor (each lasting for two minutes). From scans seven to ten, two neutral recordings (no stimulation) and two traumatic stress recordings followed by stimulation were applied. Then, the participants took a 90-minute break. After the break, four more scans were taken that included two neutral recordings and two traumatic stress recordings followed by stimulation, respectively. In short, the first day included audible delivery of six neutral scripts, six traumatic stress scripts followed by stimulation, and two stimulations without acute stress in 14 HR-PET scans. All neutral/traumatic recordings were approximately one minute in duration. The second and third days were the same as each other: they did not include brain imaging and they focused on non-personalized mental stressors. Baseline signals were recorded from the participants, and they underwent a public speech and a mental arithmetic task, both immediately followed by stimulation. In the public speech task, the participants were given two minutes preparation time to prepare a defense statement in a scenario they were accused of theft at a shopping mall. Their speech was immediately followed by stimulation. Following eight minutes in silence after stimulation stopped, the participants were required to answer a series of arithmetic questions as fast as possible for three minutes. Immediately after the arithmetic task, another stimulation was applied, and the participants waited for eight more minutes in silence (post-stimulation period). For both mental stress tasks, negative feedback was provided for incorrect answers and delayed response times to exaggerate the stress effect. After this mental stress paired with stimulation paradigm, a 90-minute break was given. After the break, a third stimulation was applied without any acute stressor.

### Transcutaneous Cervical Vagal Nerve Stimulation & Blinding

Both active tcVNS and sham stimuli were administered using hand-held devices (GammaCore, ElectroCore, Basking Ridge, New Jersey) with identical appearance, placement, and operation. The researcher identified the carotid pulsation on the left neck, and collar electrodes were placed at this location, using conductive electrode gel (Gammacore, Electrocore, Basking Ridge, New Jersey). Figure S3b shows tcVNS electrode placement. Active tcVNS produces an AC voltage signal consisting of five 5 kHz sine waves, repeating at a rate of 25 Hz (once every 40 milliseconds). The sham produces a biphasic, stepped voltage signal consisting of 0.2Hz pulses (once every five seconds). The peak voltage amplitudes for active and sham device are 30V and 14V, respectively. Figure S4 presents the active and sham stimulus waveforms. Throughout the protocol, the researcher gradually increased the stimulation intensity with a roll switch to the maximum the participant can tolerate, without pain. The active group received 20.3 V (± 7.5 SD), and sham group received 13.6V (± 1.4 SD) averaged across all uses over three days. No participants reported lack of sensation. An active stimulation amplitude higher than 15V using the studied device was previously reported to create vagal somatosensory evoked potentials associated with vagal afferent activation reliably, that are also activated with VNS implants (50).

High frequency voltage signals (such as the active stimulus) pass through the skin with minimal power dissipation due to the low skin-electrode impedance at kHz frequencies; in contrast, lower frequency signals (such as the sham stimulus) are mainly attenuated at the skin-electrode interface due to the high impedance (62). Accordingly, the active device operating at higher frequencies can deliver substantial energy to the vagus nerve to facilitate stimulation, while the voltage levels appearing at the vagus would be expected to be orders of magnitude lower for the sham device and thus vagal stimulation is highly unlikely. Nevertheless, since the sham device does deliver relatively high voltage and current levels directly to the skin, it activates skin nociceptors, causing a similar feeling to a pinch. This sensation is necessary for blinding of the participants, and is thought as a critical detail by the authors for the valuation of the potential treatment in psychiatric populations.

The randomization of the active tcVNS or sham stimulus groups were conducted with an online tool using simple randomization with group allocation completed by an individual who was dissociated from enrollment, data collection, or analysis. The devices were pre-numbered by the manufacturer who were not involved in the research design. The participants, clinical staff, and researchers were blinded to the stimulus type. Later, stimulus grouping (active or sham) was unblinded for the interpretation of statistical analysis, after data processing was completed.

### Physiological Monitoring

Physiological data were collected by the measurement of ECG, PPG, SCG, EDA, RSP, and BP, Figure S3a details the electrode placement for each participant. Wireless 3-lead ECG and piezoresistive strap-based RSP were collected through RSPEC-R amplifiers; transmissive, index finger-based PPG and inner palm-based EDA were collected through PPGED-R amplifiers from Biopac Systems (Goleta, CA). A low noise 356A32 accelerometer (PCB Electronics, Depew, NY) was taped with a Kinesio tape on mid-sternum for SCG monitoring, with Z-axis surface touching the sternum, aligning with the dorsoventral movement of the heart. For EDA measurement, an isotonic electrode gel (GEL101) and pre-gelled isotonic electrodes (EL507) were used (Biopac Systems, Goleta, CA). Continuous ECG, PPG, SCG, EDA, and RSP data were simultaneously transmitted to a 16-bit MP150 data acquisition system at 2kHz sampling rate (Biopac Systems, Goleta, CA). Non-continuous, cuff-based systolic (SBP) and diastolic blood pressure (DBP) values were recorded periodically with an Omron blood pressure cuff for baseline, stress, stimulation, and post-stimulation intervals.

### Signal Processing & Parameter Extraction

The physiological signals were processed in MATLAB (R2020a, Mathworks, Natick MA) and the following parameters were extracted: heart rate (HR), pre-ejection period (PEP), amplitude of PPG, pulse arrival time (PAT), respiration rate (RR), width (RW), respiration prominence (RP), low frequency and high frequency heart rate variability (LF HRV, HF HRV), non-linear heart rate variability (SD1, SD2), skin conductance level (63), skin conductance response (64), frequency of non-specific skin conductance responses (f_NSSCR_), and latency of skin conductance response (L_SCR_). Signal processing steps were matched to the previous work investigating non-PTSD population (57).

#### Pre-processing

ECG, PPG, SCG signals were band-pass filtered with cut-off frequencies (f_c_) 0.6-40Hz for ECG, 0.4-8Hz for PPG, and 0.6-25Hz for SCG to cancel the noise out of the bandwidth. EDA was high-pass filtered (f_c_=0.15Hz) to obtain the fast varying AC component. Its DC level was calculated from the signal’s trend. The R-peaks of ECG were detected using thresholding (i.e. *findpeaks* MATLAB function), and were used to calculate HR and HRV. PPG and SCG signals were segmented referenced to the R-peaks of ECG signal. SCG and feature points related to continuous blood pressure on PPG are prone to artifacts: SCG captures heart motion, body motion, and vibration due to talking; motion artifacts in PPG might instantaneously push or move back the pulse arrival point on the waveform to calculate PAT. To mitigate these, an additional exponential moving averaging step (65) was carried out after segmenting for these signals, as described below. The extracted parameters are summarized as follows:

#### Heart rate and heart rate variability measures

Frequency-domain analysis and joint time-frequency analysis (Poincaré Method) were used to extract the autonomic measures LF HRV, HF HRV, LF/HF HRV, SD1, SD2, SD1/SD2 values. Based on previous studies, the LF HRV and SD2 are a reflection of baroreflex sensitivity (BRS) (70). Individuals with PTSD are reported to have reduced BRS in previous studies (16), therefore these biomarkers of BRS have been computed. As low-frequency HRV measures require at least five minutes of continuous ECG signal, the comparisons from the start to the end of that days (during rest periods) were the focus of our analyses using a MATLAB toolbox that has been reliably validated with different datasets before (71).

#### Pre-ejection period

PEP is determined by the ventricular electromechanical delay of isovolumic contraction period. As a cardiac timing interval inversely related to cardiac contractility, decreased PEP reflects increased cardiac ß_1_ receptor stimulation and cardiac sympathetic activity, also termed as “effort” in the literature (67, 72, 73). It can be obtained from simultaneously collected ECG (reference for electrical depolarization of the heart) and SCG (reference for aortic valve opening of the heart) signals by calculating the time delay from ECG R-peaks to the aortic opening point on SCG beats with a three-beat exponential moving averaging paradigm (65, 68).

A decrease in PEP denotes increased cardiac contractility and sympathetic activity.

#### PPG amplitude and pulse arrival times

Calculated the beat-by-beat amplitude of the peripheral blood volume pulse, photoplethysmogram (PPG) amplitude is a peripheral sympathetic activity index, reflecting the dilation or constriction of peripheral blood vessels. An increase in PPG amplitude suggests vasodilation at the local area where the signal is acquired from (index finger). PAT is the time delay between the electrical stimulation of the heart (obtained from ECG R-peaks) to the “foot” (or trough) of a distal arterial waveform (taken from the index finger). It is the sum of the pulse transit time (PTT), which is inversely related to blood pressure, and the PEP (74). PPG amplitude and PAT are vascular-dominated measures among other factors (75, 76). PAT values were calculated with a five-beat exponential moving averaging.

#### Respiratory measures

Due to the role of the vagus nerve in efferent parasympathetic activity, respiratory measures that take part in the regulation of parasympathetic activity were extracted: respiratory rate (RR), width (RW), prominence (RP). As the RSP signal was taken with a piezoresistive strap, it had varying DC offset due to tightening/loosening of the strap. To detrend the signal, a sixth order polynomial was fit to the signal at each interval of interest, and points representing inhalation and exhalation were located. RR was calculated as the instantaneous rate of peak appearance, RW was calculated as the width of each respiration cycle, and RP was calculated as the prominence of each respiration peak.

#### Electrodermal activity measures

Skin conductance level (SCL) and its slope, skin conductance response, frequency of non-specific peaks (f_NSSCR_), and latency of SCR (L_SCR_) were extracted as sweat gland activity measures (66, 77).

#### Blood pressure measures

Periodic values of SBP and DBP were used to find pulse pressure (PP) and mean arterial pressure (MAP) using a clinical grade upper arm BP cuff (Omron Healthcare, Kyoto, Japan).

### Statistical Analysis

The baseline characteristics between the device groups were compared in Table S2. For this comparison, normality was assessed using Shapiro-Wilk test. Student’s t-test, Wilcoxon rank-sum tests, and chi-squared tests were used for normal continuous, non-normal continuous, and categorical variables, respectively. Stimulation-only administrations (n = 2 on day one, n = 1 on days two and three) were expressed as change values (from baseline) for stimulation and post-stimulation intervals and subsequently averaged across days. For stress tasks, data during stress, stimulation, and post-stimulation intervals were extracted. Data were then averaged across stress types (six traumatic, two public speech, two mental arithmetic stressors) and change values (from baseline) of intervals (stress, stimulation, post-stimulation) were computed. For mental tasks that require speaking (public speech, mental arithmetic), stress data were extracted during times without vocalizations (immediately before task) to avoid unwanted signal noise while talking. Longer intervals of ECG (exceeding three minutes), as extracted as baseline, following a break, and end of day were used to assess HRV changes. Four essential comparisons were performed to assess differences between the device groups: stimulation without stress, stimulation following traumatic stress, public speech stress, mental arithmetic stress. Data in bar plots represent raw (unadjusted) mean ± 95% confidence interval (95% CI). Mixed models with repeated measures included random effect for each participant, used an unstructured correlation matrix, and were adjusted for age. The beta coefficients (ß) from the mixed models indicate the adjusted average percent or absolute differences in active group compared to sham group. ß were reported along with adjusted 95% CI, p-values, and effect sizes (d, based on Cohen’s d for independent observations)(78) in results and figure captions in ß (±CI, d, p) format. A two-sided p<0.05 denoted statistical significance. All statistical analyses were performed using SAS 9.4 (SAS Institute, Cary, NC) and MATLAB (R2020a, Natick, MA).

## Results

### tcVNS consistently decreases SNS in absence of stress over multiple days

Figure 1 presents the raw values for autonomic (SD1/SD2), cardiovascular (HR), and vascular (PPG Amplitude and PAT) tone. Compared to sham, active tcVNS increased SD1/SD2 (Figure 1A, post-stimulation, ß (±CI, d, p) 14.1% (±11.6%, d=0.43, p=0.019)), decreased HR (Figure 1B, following stimulation, 2.7% (±2.0%, d=0.21, p=0.009)), increased PPG amplitude (inversely associated with peripheral sympathetic activity, Figure 1C, during stimulation, 43.4% (±43.4%, d=0.53, p=0.049); following stimulation 73.1% (±63.2%, d=0.7, p=0.025)), and increased PAT (inversely associated with peripheral sympathetic activity, Figure 1D, during stimulation, 2.5% (±2.2%, d=0.26, p=0.026)). The results were similar when the days were evaluated separately (Figures S5 and S6).

**Figure 1.**
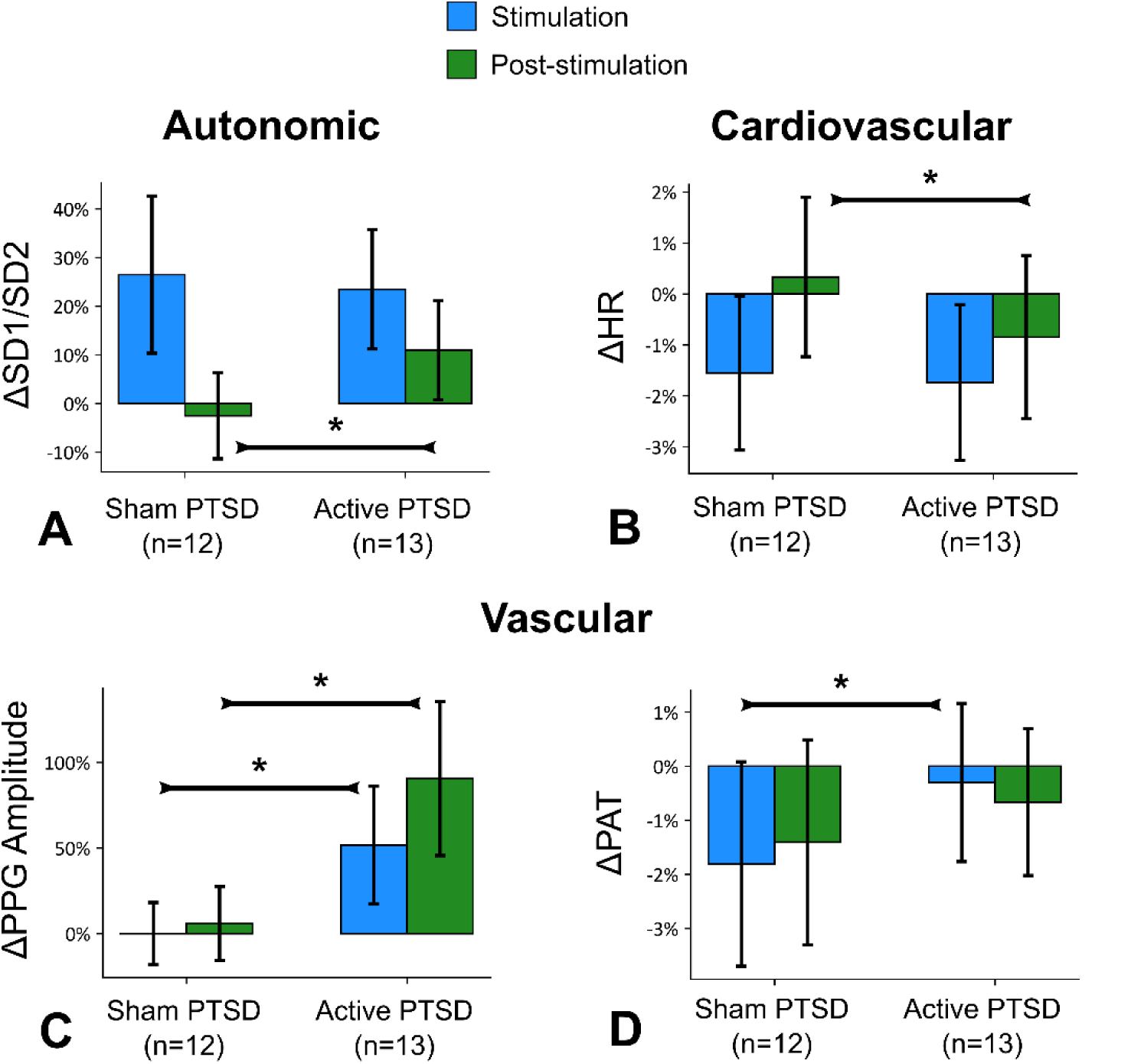
tcVNS without acute stress. Outcomes for stimulation without acute stress, merged from all days. Bars represent the unadjusted mean changes from baseline, error bars: 95% CI, values calculated from raw data, * indicates p<0.05. ß coefficients, adjusted CI, effect sizes (d), and p-values were reported in ß (±CI, d, p) format. Active tcVNS group experienced the following relative to sham after adjustments: **(A)** The ratio of short-term variability to long-term variability (SD1/SD2) increased following stimulation by 14.1% (±11.6%, d=0.43, p=0.019). **(B)** HR decreased following stimulation by 2.7% (±2.0%, d=0.21, p=0.009). **(C)** PPG amplitude increased during stimulation by 43.4% (±43.4%, d=0.53, p=0.049) and following stimulation by 73.1% (±63.2%, d=0.67, p=0.025). **(D)** PAT increased during stimulation by 2.5% (±2.2%, d=0.26, p=0.026).

### tcVNS reduces sympathetic tone following exposure to personalized traumatic scripts

Stimulation following exposure to personalized traumatic scripts resulted in marked changes in autonomic reactivity, similarly to stimulation without stress. Figures 2A-C illustrate changes in from the baseline state for the three intervals: traumatic stress, stimulation, and post-stimulation, merged from all six traumatic stressors. Relative to sham, active tcVNS decreased HR (Figure 2A, during stimulation, 5.6% (±3.6%, d=0.43, p=0.003); following stimulation, 3.9% (±3.0%, d=0.29, p=0.013)), increased PPG amplitude (Figure 2B, during stimulation, 30.8% (±28.0%, d=0.41, p=0.032)), increased PAT (Figure 2C, during combined traumatic stress, 9.2% (±3.0%, d=0.15, p<0.0001); during stimulation, 2.2% (±2.2%, d=0.42, p=0.045); following stimulation, 6.2% (±1.9%, d=0.57, p<0.0001)), indicating attenuation in the elevated autonomic tone due to stress. These effects were not initially observed, as no differences (p > 0.05) were found between active and sham during the first traumatic script. Figure S7D-F presents the stress reactivity across each traumatic script for these biomarkers. tcVNS also decreased long-term heart rate variability after multiple traumatic stress and stimulation protocol: Following the repeated traumatic stress protocol, tcVNS decreased SD2 obtained from Poincaré plot (31.7ms (±15.5ms, d=1.02, p=0.0002); Figure 3).

**Figure 2.**
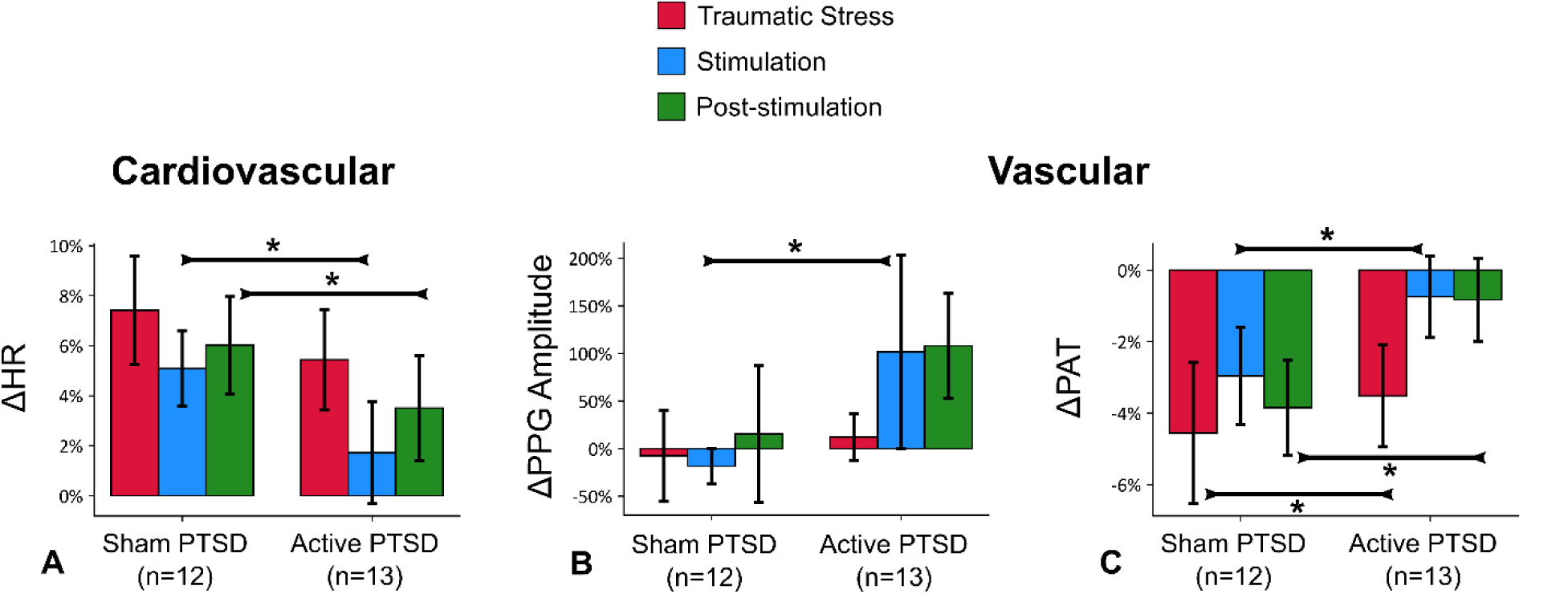
tcVNS after traumatic stress. Outcomes for stimulation following traumatic stress (all six scripts). Bars represent the unadjusted mean changes from baseline, error bars: 95% CI, values calculated from raw data, * indicates p<0.05. ß coefficients, adjusted CI, effect sizes (d), and p-values were reported in ß (±CI, d, p) format. Active tcVNS group experienced the following relative to sham after traumatic stress after adjustments: **(A)** HR decreased during stimulation by 5.6% (±3.6%, d=0.43, p=0.003), and following stimulation by 3.9% (±3.0%, d=0.29, p=0.013). **(B)** PPG amplitude increased during stimulation by 30.8% (±28.0%, d=0.41, p=0.032). **(C)** PAT decreased less during traumatic stress by 9.2% (±3.0%, d=0.15, p<0.0001), stimulation by 2.2% (±2.2%, d=0.42, p=0.045), and following stimulation by 6.2% (±1.9%, d=0.57, p<0.0001).

**Figure 3.**
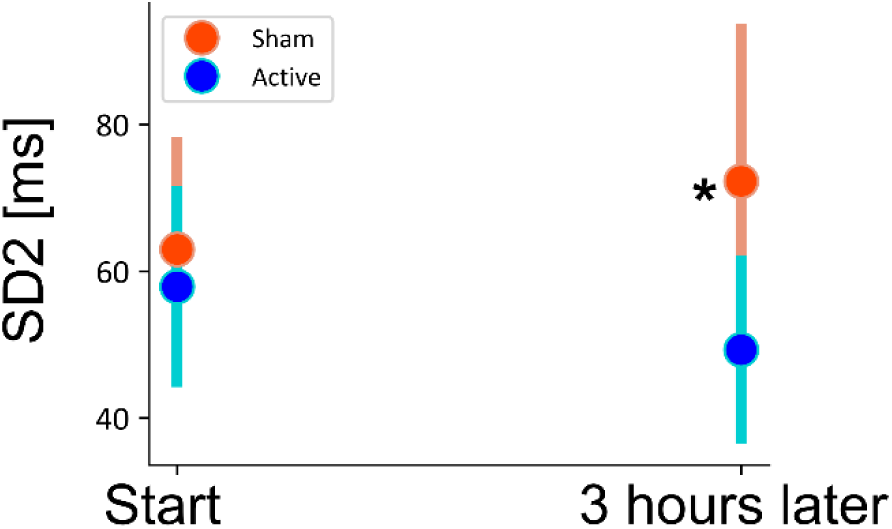
Change in long-term heart rate variability (SD2) for multiple stimulation protocol following traumatic stress (four traumatic stress and six stimulation administrations on the first day). Bars represent the unadjusted mean changes from baseline, error bars: 95% CI, values calculated from raw data, * indicates p<0.05. ß coefficients, adjusted CI, effect sizes (d), and p-values were reported in ß (±CI, d, p) format. Active tcVNS group experienced decrease in SD2 after the multiple stress protocol by 31.7ms (± 15.5ms, d=1.02, p=0.0002) after adjustments.

### tcVNS affects cardiac contractility and heart rate variability following mental stress

Figures 4A-D summarize the effects of tcVNS when applied after two different mental stress tasks: public speech and mental arithmetic on the second and third days. SD1/SD2 increased during stimulation right after speech task (Figure 4A, inversely related to sympathetic activity, during stimulation, 23.1% (±21.1%, d=0.71, p=0.033)) and after mental arithmetic test (Figure 4B, during stimulation, 41.2% (±22.5%, d=0.44, p=0.001)).

**Figure 4.**
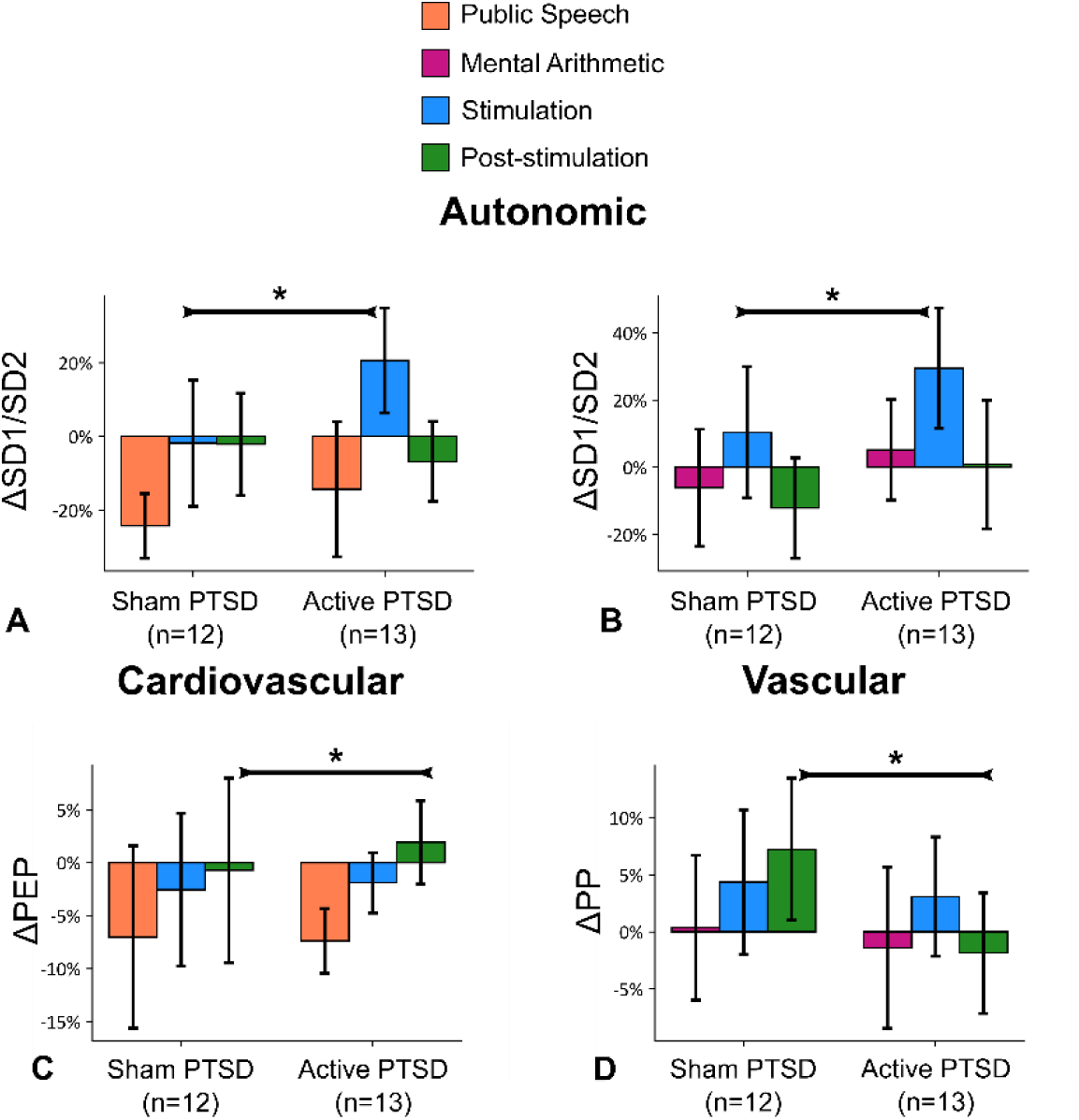
tcVNS after mental stress. Outcomes for stimulation following two types of mental stress, public speech and mental arithmetic. Bars represent the unadjusted mean changes from baseline, error bars: 95% CI, values calculated from raw data, * indicates p<0.05. ß coefficients, adjusted CI, effect sizes (d), and p-values were reported in ß (±CI, d, p) format. Active tcVNS group experienced the following relative to sham after adjustments: **(A)** SD1/SD2 increased during stimulation right after speech task by 23.1% (±21.1%, d=0.71, p=0.033). **(B)** Similar to the speech task, SD1/SD2 increased by 41.2% (±22.5%, d=0.44, p=0.001). **(C)** PEP increased following stimulation after speech task by 6.8% (±5%, d=0.16, p=0.009). **(D)** PP decreased following stimulation after mental arithmetic by 9.6% (±9.7%, d=0.68, p=0.049).

Active tcVNS increased PEP (Figure 4C, inversely related to cardiac sympathetic activity) following stimulation compared to sham (6.8% (±5%, d=0.16, p=0.009)), indicating a decrease in cardiac contractility and sympathetic activity. Active tcVNS also decreased PP following stimulation after the mental arithmetic task compared to sham (9.6% (±9.7%, d=0.68, p=0.049); Figure 3D), indicating a decrease in vascular reactivity

## Discussion

This study showed that tcVNS modulates autonomic, cardiovascular, and vascular measures in PTSD with or without exposure to traumatic and mental stress. The broad interpretation of the changes due to tcVNS are similar to the study involving non-PTSD controls (57) (i.e., reduction of sympathetic tone at baseline and blocking sympathetic responses to stress). Additionally, the effects of tcVNS on vascular measures (i.e., PPG amplitude) persist as markers of autonomic changes with tcVNS independent of disease status.

Active tcVNS decreased sympathetic arousal as measured by autonomic, cardiovascular, and vascular measures across multiple days and types of stressors. Results were seen on the first day after multiple exposures to personalized traumatic script stress, and on the afternoons of the second and third days after exposure to mental stress challenges (public speaking, mental arithmetic) in the morning (Figures S5–S6). PPG amplitude was a persistent biomarker of stimulation regardless of the disease status. We found greater SD1/SD2 response to tcVNS than sham, while the other frequency domain metrics were not affected by tcVNS. While the biological significance of this measure/finding is not clear, it is complemented by auricular VNS studies in which frequency-domain HRV improved (29, 30, 51). SD1/SD2 is a non-linear Poincaré-based HRV, which is less studied in literature, although another study noted that it is negatively associated with diabetes (79). Hemodynamic measures (BP, HR) have been studied more, and been mixed throughout tcVNS studies: tcVNS decreased HR in humans (51) and rats, albeit only momentarily (55). Other studies, however, have not observed any autonomic or cardiovascular changes (56).

tcVNS improved recovery from traumatic stress (reduced HR, increased PAT) and decreased peripheral sympathetic activity (decreased PPG amplitude). The results except PPG amplitude (HR and PAT) differ from the healthy-cohort results for traumatic stress. Additionally, no difference in PEP or EDA were noted for the PTSD cohort. PAT reactivity to traumatic stress was comparable between the groups when data from only the first script was analyzed (Figure S7A-C). With the merged data (Figure 2C), sham group experienced more reactivity to stress. Patients with PTSD fail to habituate to repeated exposure of stress (80, 81). The therapeutic potential of tcVNS is shown by its effect in decreasing stress reactivity. These results suggest that repeated tcVNS enhances resilience in the face of repeated stress in patients with PTSD.

An interesting outcome was the decrease in SD2 in the active tcVNS group with PTSD (Figure 3), which might indicate increased BRS. Reduced BRS is associated with increased mortality, higher inflammation (82), increased depressive symptoms (83), and increased risk of myocardial infarction (84, 85), and lower BRS has been proposed as a risk stratification for multiple cardiac mortality conditions (86, 87). It is also known that patients with PTSD have impaired autonomic modulation and autonomic inflexibility as measured by frequency-domain HRV (16). We observed dampened SD2, which may also support this concept regarding BRS as an important moderating factor (29).

Active tcVNS improved cardiac contractility recovery (PEP) following the speech task. PEP is an index of effort-related cardiac activity (72), with greater PEP indicated decreased effort and cardiac sympathetic activation. PEP is responsive to tasks requiring effortful active coping (72), similar to the speech task, potentially indicating tcVNS mitigates effort during the speech task. PEP has been shown previously to respond differently to challenge or threat conditions, specifically resulting in decreases with challenge and minimal changes with threat (88). In the current study, PEP decreased with the speech task in both groups, and the decrease was mitigated with active tcVNS. The traumatic stress perhaps could be regarded as threat for patients with PTSD. PEP outcomes for speech tasks and (the lack of) PEP outcomes for traumatic stress might be due to the perceptional differences for challenge versus threat (89). A comparison of previous works in PTSD notes more than double cortisol release with cognitive challenge (90), compared to the cortisol levels with traumatic stress (91). Although there is no direct statistical comparison between these studies, the magnitude differences in cortisol levels are apparent.

Several aspects of this study might limit the generalization of the results. The active group was female-dominated due to the small sample size. However it is important to note that PTSD is twice more prevalent in females than males (92). The timing of the stimulation is also relevant to consider in the interpretation of our results. Prior animal and human studies initiated the stimulation before or during the stimuli (25, 46, 93–95). In our study, we stimulated immediately at the end of the exposure to stress in terms of listening to scripts or performing mental stress (16, 17). Given instructions to hold images of trauma in the mind, and based on our prior work showing that physiological indicators of stress persist after termination of the task, we feel that this timing corresponds to the peak period of stress as experienced by the individual. Our findings on the effects of tcVNS using this protocol on autonomic function, however, are consistent with prior preclinical and clinical studies of VNS using various stimulation timings and protocols (24, 25, 29, 46, 96). Future studies should compare stimulation before, during, and at the termination of stress protocols.

In summary, using a multimodal sensing approach, we found that tcVNS at rest and paired with various stressors modulates the autonomic nervous system and cardiovascular reactivity. We demonstrated feasibility of use of wearable sensing devices for measurement of novel physiological markers in patients with PTSD. These modalities can be used in a home setting to assess target engagement and treatment efficacy for personalized neuromodulation. Future work should focus on methods to evaluate longitudinal outcomes (97) and parameter determination studies (28) for selective modulation of autonomic tone and to utilize these modalities in assessment of response to neuromodulation treatments in PTSD.

## Data Availability

The data and MATLAB/SAS codes, that support the findings of this study, are available from the corresponding author (Nil Gurel) upon reasonable request.

## SUPPLEMENTARY FIGURES

**Figure S1.**
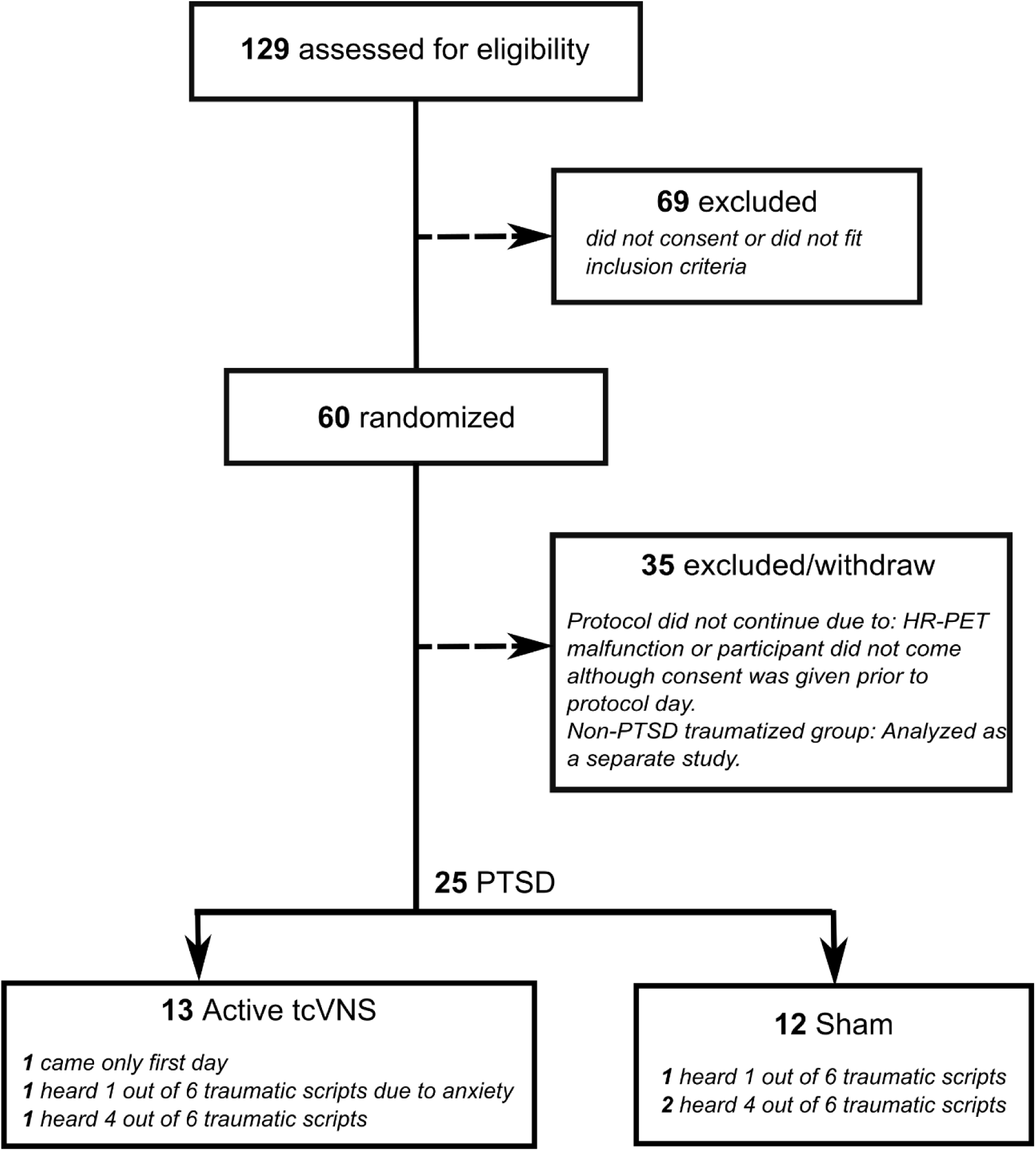
Consolidated Standards of Reporting Trials (CONSORT) diagram of the study.

**Figure S2.**
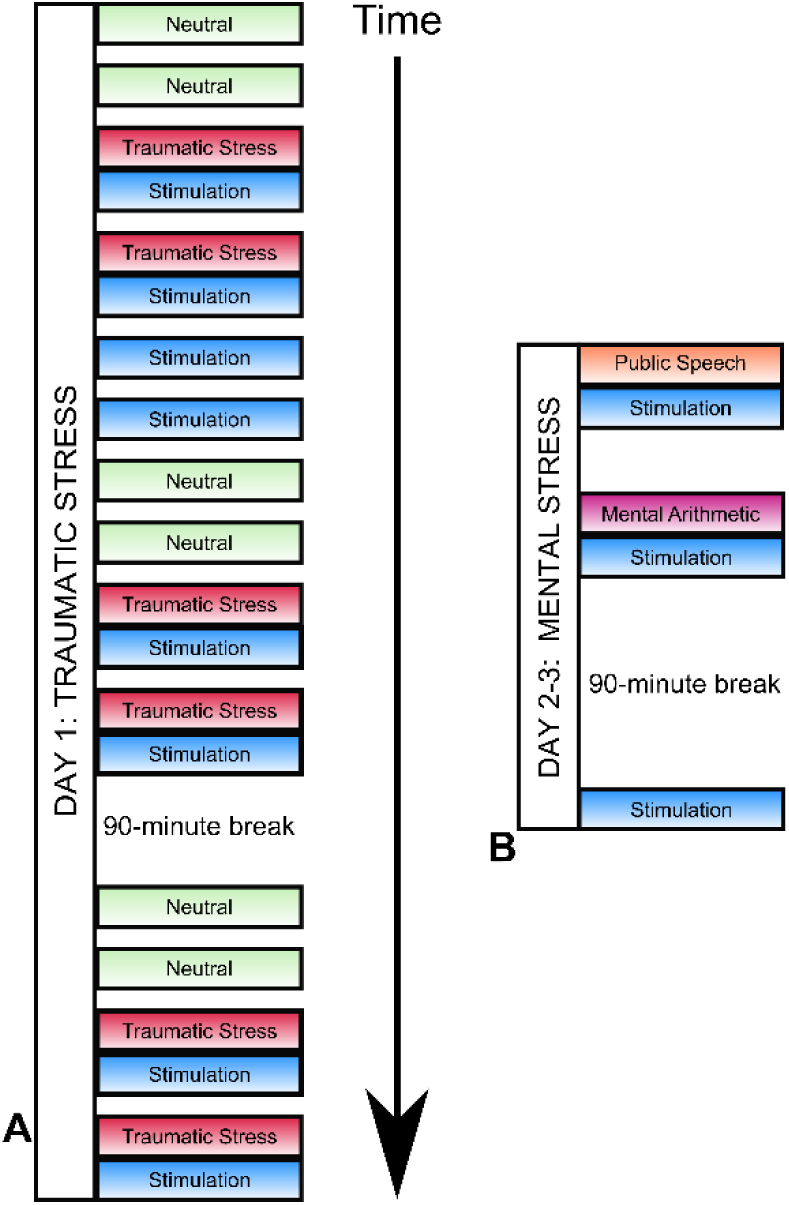
Protocol description. **(A)** The first day included traumatic stress through headphones. After each traumatic stress prompt, stimulation (active or sham) was applied immediately. **(B)** Second and third days included two types of mental stress, public speech and mental arithmetic. After each stressor, stimulation was applied immediately. After a 90-minute break from the mental stress protocol, participants received stimulation without acute stress.

**Figure S3.**
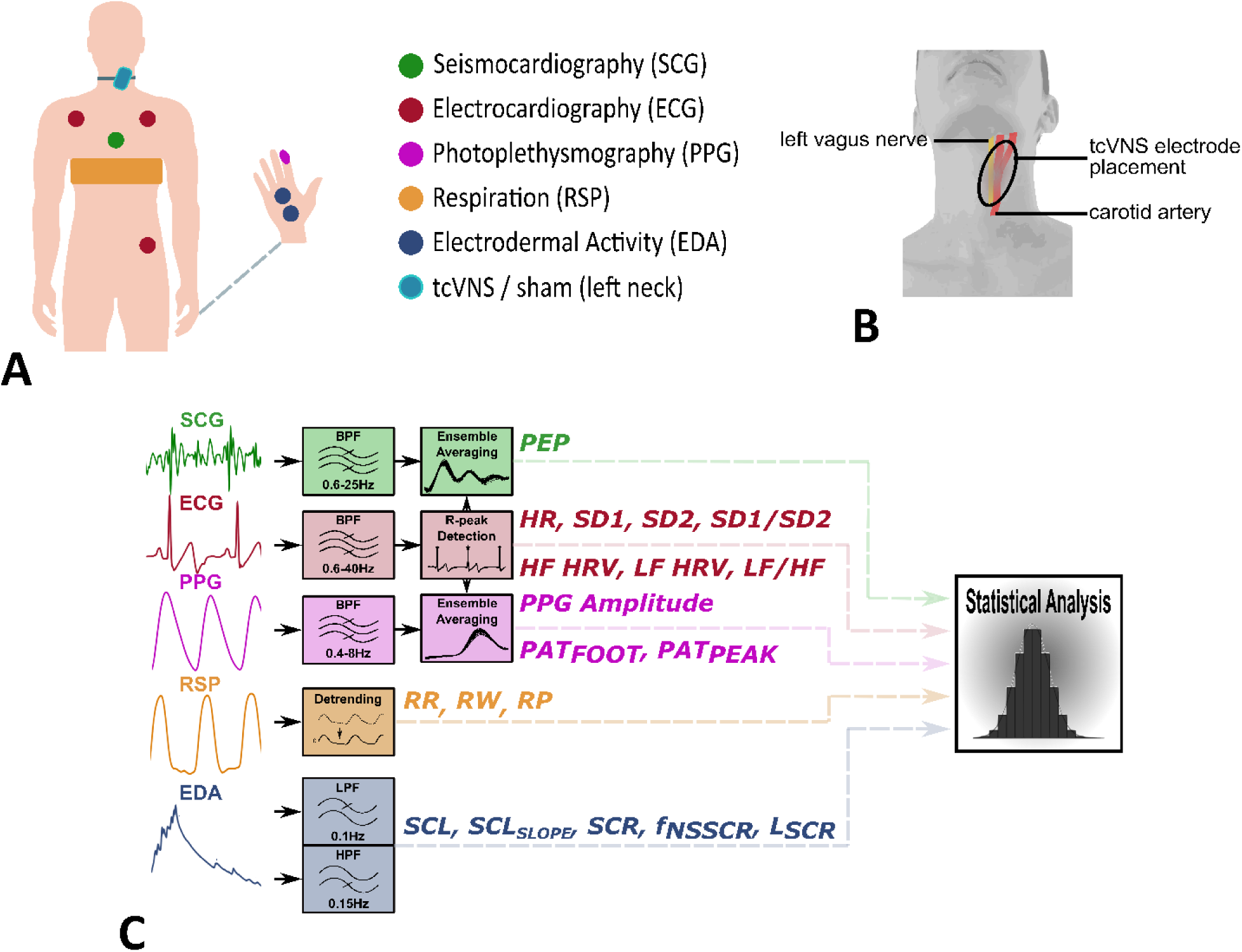
Data collection and signal processing summary. **(A)** Non-invasive sensing modalities shown on participant, active or sham stimulation was applied from left neck. **(B)** Representation of relative locations of left carotid arteries and left vagus nerve. tcVNS electrodes were placed onto the area where the carotid pulsation was located. **(C)** Signal processing and feature extraction summary.

**Figure S4.**
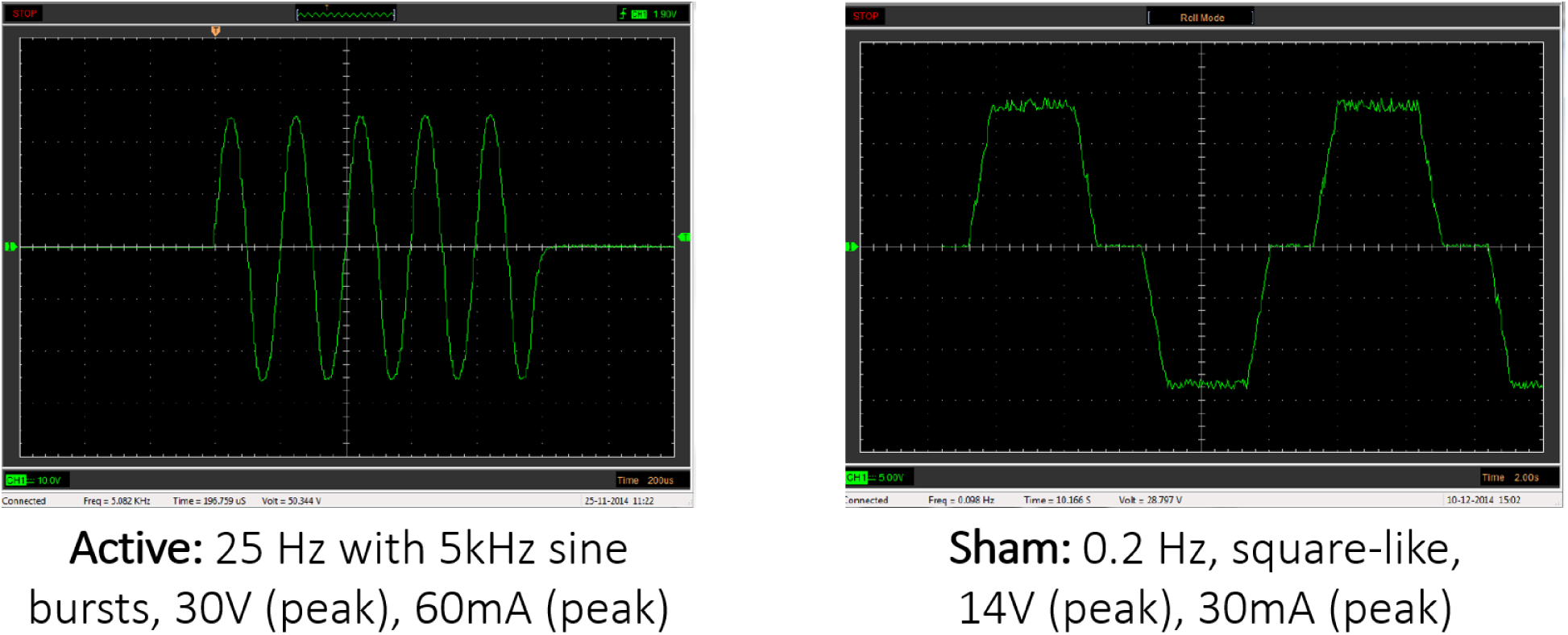
Active tcVNS (left) and sham (right) waveform characteristics as plotted from oscilloscope. The active tcVNS produces an AC electrical signal consisting of five 5,000 Hz pulses (200 microseconds each), repeating at a rate of 25 Hz (once every 40 milliseconds), at 30V (peak) amplitude. The waveform of active pulse is approximately a sine wave. The sham produces an electrical signal consisting of 0.2 Hz pulses, repeating at a rate of every 5 seconds. The waveform of the sham consists of 2 seconds at +14V (peak) then 1 second at 0 V then 2 seconds at −14V (peak) approximately in a stepped wave pattern. Both devices have identical placement and operation, both devices stop automatically after 2 minutes. The researcher increases the stimulation intensity to the maximum the participant can tolerate, without pain, regardless of the device type.

**Figure S5.**
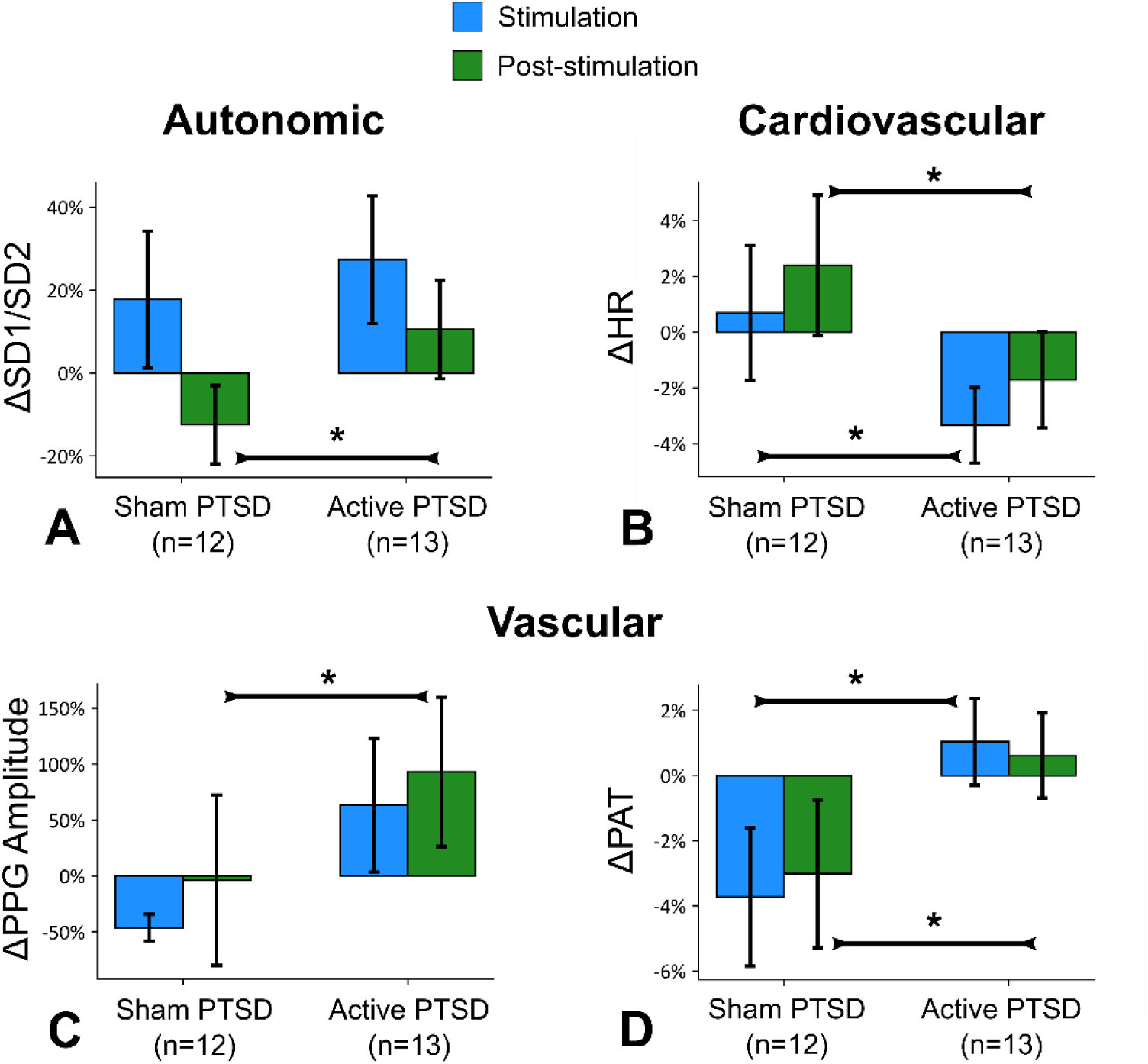
tcVNS without acute stress, data from the first day. Bars represent the unadjusted mean changes from baseline, error bars: 95% CI, values calculated from raw data, * indicates p<0.05. ß coefficients, adjusted CI, effect sizes (d), and p-values were reported in ß (±CI, d, p) format. Active tcVNS group experienced the following relative to sham after adjustments: **(A)** Increase in SD1/SD2 following stimulation by 21.9% (±21.9%, d=0.90, p=0.048). **(B)** Decrease in HR during stimulation by 3.9% (±3.7%, d=0.85, p=0.023), and following stimulation by 4.3% (±4.2%, d=0.78, p=0.035). **(C)** PPG amplitude increased following stimulation by 121.2% (±112.5%, d=0.54, p=0.047). **(D)** PAT increased during stimulation by 4.9% (±3.1%, d= 1.10, p=0.003), and following stimulation by 3.8% (±3.4%, d=0.81, p=0.023).

**Figure S6.**
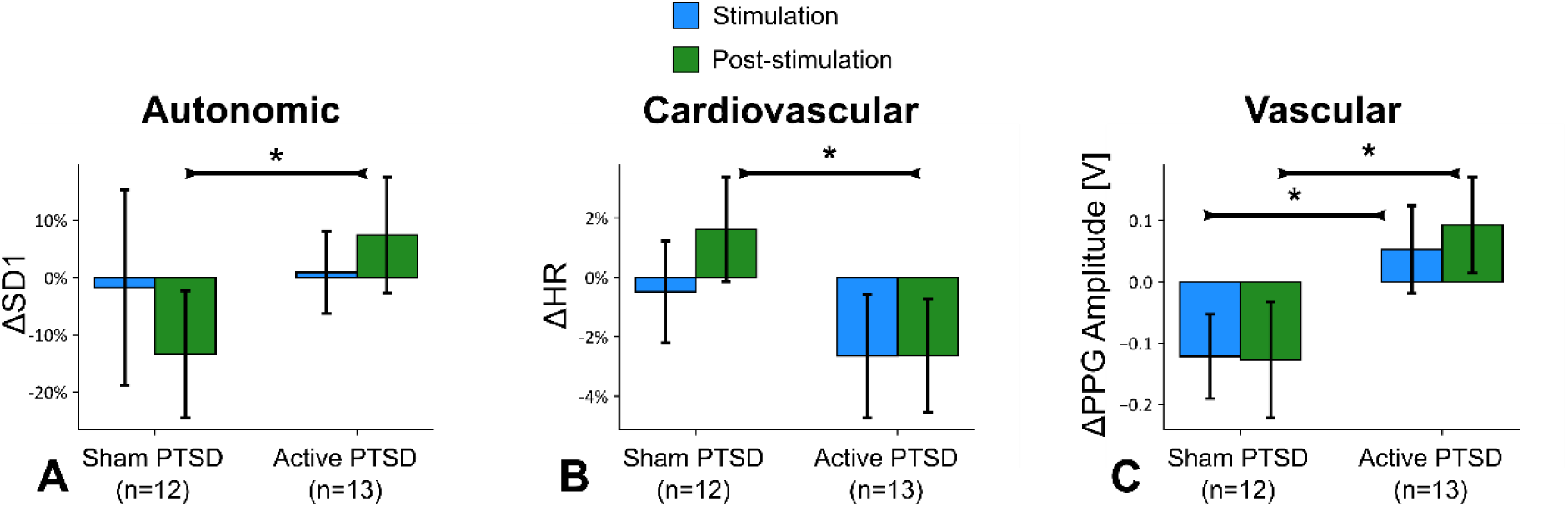
tcVNS without acute stress, data from the second and third days. Bars represent the unadjusted mean changes from baseline, error bars: 95% CI, values calculated from raw data, * indicates p<0.05. ß coefficients, adjusted CI, effect sizes (d), and p-values were reported in ß (±CI, d, p) format. Active tcVNS group experienced the following relative to sham after adjustments: **(A)** Short-term-variability (SD1) increased following stimulation by 20.3% (±16.5%, d=0.84, p=0.018) **(C)** HR decreased following stimulation by 4% (±2.9%, d=0.46, p=0.007). **(C)** PPG amplitude increased during stimulation by 0.18a.u. (±0.1a.u., d= 1.01, p=0.004), and following stimulation by 0.23a.u. (±0.1a.u., d=1.03, p=0.003).

**Figure S7.**
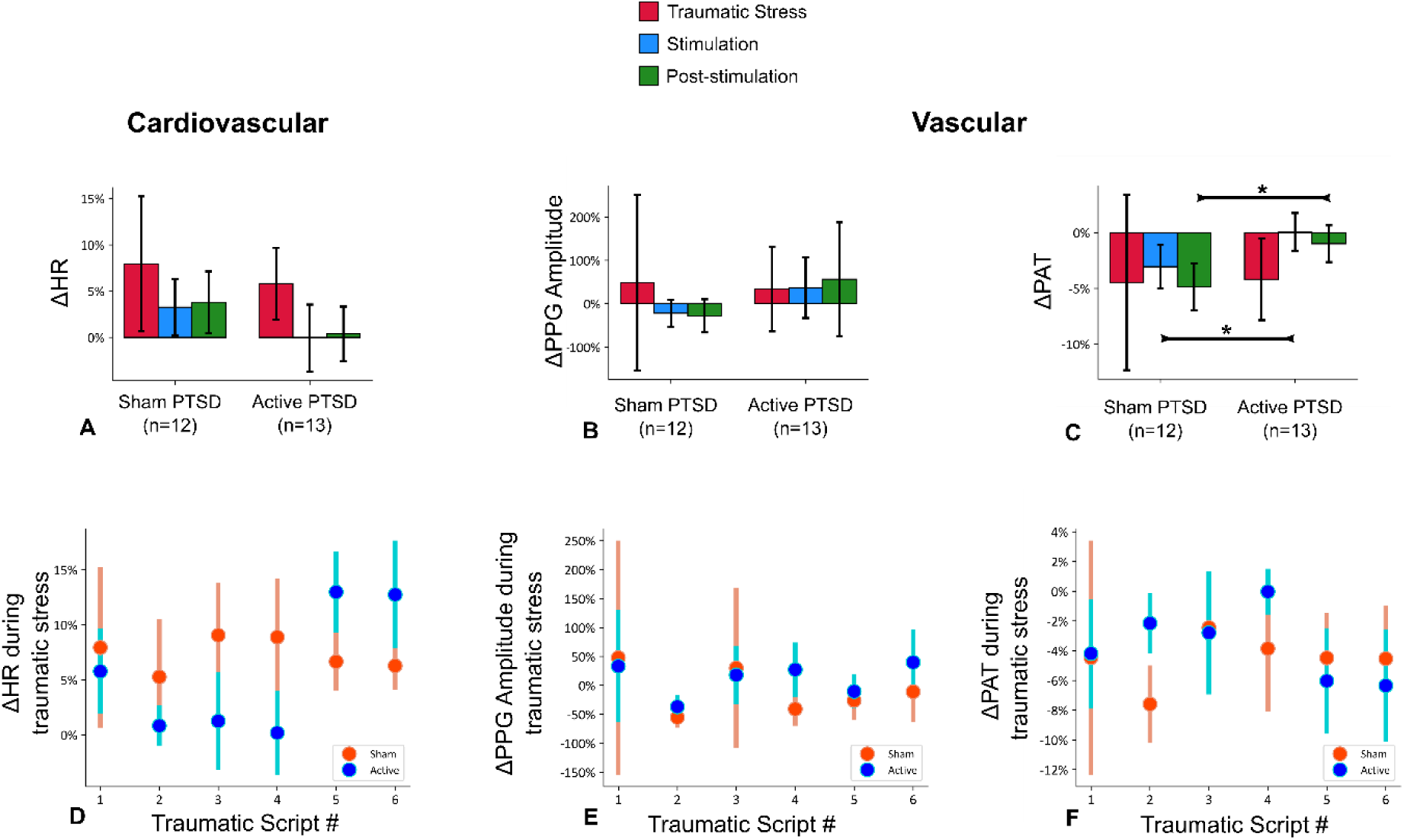
Unadjusted mean changes ± 95% confidence interval for the first traumatic stress script and the change in traumatic stress reactivity (or habituation) from the first to the sixth script between the groups (D-F). * indicates p<0.05. ß coefficients, adjusted CI, effect sizes (d), and p-values were reported in ß (±CI, d, p) format. **(A)** No significant difference in HR between groups in any of the intervals. **(B)** No significant difference in PPG amplitude between groups in any of the intervals. **(C)** PAT increased during stimulation by 3.3% (±2.9%, d=0.95, p=0.027), and following stimulation by 4.2% (±2.9%, d= 1.15, p=0.005) after adjustments indicating attenuation in the elevated autonomic tone due to stress. **(D)** HR reactivity to traumatic stress scripts as the protocol transitions from the first to the sixth script. **(E)** PPG amplitude reactivity. **(F)** PAT reactivity.

## SUPPLEMENTARY TABLES

**Table. S1.**
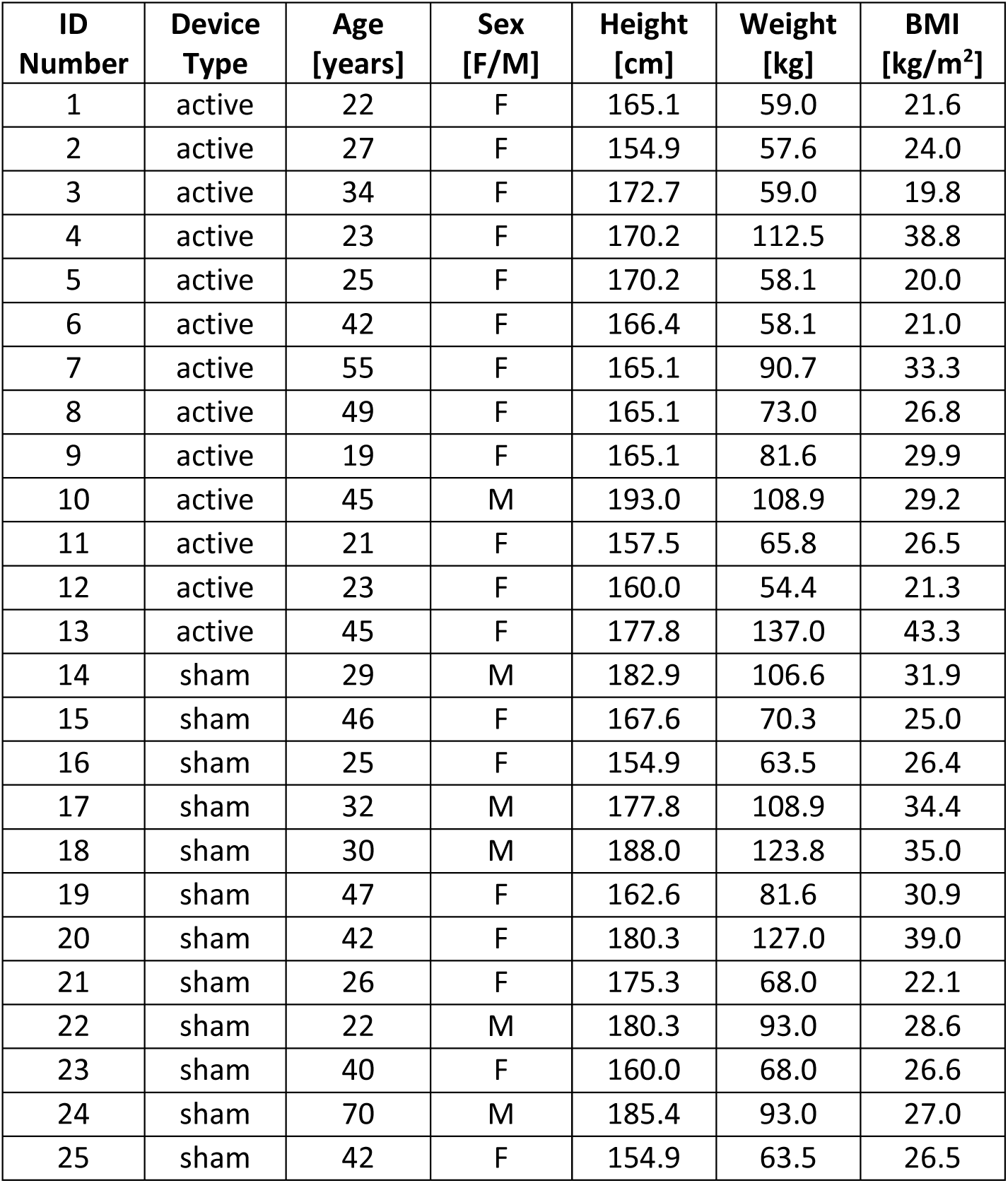
Anthropometric information per participant. F, female; M, male; BMI, body mass index.

**Table. S2.** Anthropometric and physiological parameter information per device group. P: p-value for the comparison between groups.

| Parameter | Active | Sham | P |
| --- | --- | --- | --- |
| Age [years] | 33.1 (12.0) | 37.6 (12.8) | 0.21 |
| Sex [F, %] | 12F, 92.3% | 7F, 58.3% | 0.06 |
| Height [cm] | 167.9 (9.3) | 172.5 (11.4) | 0.31 |
| Weight [kg] | 78.1 (25.5) | 88.9 (22.3) | 0.13 |
| BMI [kg/m<sup>2</sup>] | 27.3 (7.1) | 29.5 (4.7) | 0.42 |
| HR [bpm] | 74.4 (10.9) | 68.8 (11.0) | 0.23 |
| PEP [ms] | 77.5 (55.0) | 80.6 (34.5) | 0.87 |
| PPG [V] | 0.3 (0.3) | 0.3 (0.3) | 0.57 |
| RR [rpm] | 16.9 (3.8) | 17.5 (4.5) | 0.71 |
| RW [s] | 2.0 (0.6) | 1.8 (0.5) | 0.32 |
| RP [V] | 0.7 (0.8) | 0.7 (0.9) | 0.72 |
| PAT [ms] | 264.2 (15.1) | 272.8 (11.6) | 0.14 |
| HF HRV [ms<sup>2</sup>] | 611.7 (532.0) | 879.9 (668.3) | 0.35 |
| LF HRV [ms<sup>2</sup>] | 738.4 (645.2) | 799.5 (529.9) | 0.60 |
| LF/HF [n.u.] | 1.7 (1.4) | 1.2 (0.8) | 0.58 |
| SD1 [ms] | 22.7 (15.0) | 27.0 (13.0) | 0.48 |
| SD2 [ms] | 57.9 (25.2) | 64.7 (22.0) | 0.51 |
| SD1/SD2 [a.u.] | 378.9 (160.0) | 405.2 (113.7) | 0.67 |
| SBP [mmHg] | 126.8 (22.2) | 130.4 (17.1) | 0.57 |
| DBP [mmHg] | 77.5 (13.6) | 74.8 (9.1) | 0.79 |
| PP [mmHg] | 49.4 (10.8) | 55.7 (12.0) | 0.25 |
| SCL<sub>MEAN</sub> [μS] | 2.9 (2.9) | 4.3 (6.1) | 0.50 |
| SCL<sub>SLOPE</sub> [μS/s] | 0.3 (0.7) | 3.5 (8.5) | 0.12 |
| f<sub>NSSCR</sub> [pks/s] | 0.4 (0.3) | 0.4 (0.3) | 0.97 |
| L<sub>SCR</sub> [s] | 15.5 (8.7) | 24.1 (19.3) | 0.62 |

